# CORRELATIVE STUDY ON BODY MASS INDEX AND BLOOD PRESSURE IN THE UNITED KINGDOM: A SYSTEMATIC REVIEW OF CURRENT EVIDENCE

**DOI:** 10.1101/2023.10.13.23297003

**Authors:** David Chinaecherem Innocent, Advait Vasavada, Ramesh Kumar, Rupesh Andani, Cosmas Nnadozie Ezejindu, Mohammad Arham Siddiq, Rejoicing Chijindum Innocent, Ihuoma Chimdimma Dike, Mohamed Lounis

## Abstract

**Background:** A few studies have found a connection between body mass index (BMI) and blood pressure (BP), which may contribute to people’s health issues. A person who has a BMI greater than what is considered healthy for their height to weight ratio is more likely to have high blood pressure, which increases their risk for conditions including type 2 diabetes, gallstones, respiratory issues, and some types of cancer. Understanding the relationship between blood pressure and body mass index is crucial.

**Aim:** The overall goal of this review is to provide evidence on a correlative study of blood pressure and BMI in the United Kingdom.

**Methods:** A literature search was conducted on PsycINFO, PubMed, Web of Science, Science Direct, and Cochrane Library to identify studies addressing the primary research question. The participants for this study were individuals in the United Kingdom aged 18 years and above. The study considered studies published from 2000–2022 and quantitative studies as well as mixed-method studies. The critical appraisal risk of bias tool was used to determine the quality assessment of the studies included in this systematic review.

**Results:** 27,322 participants were involved from a total of seven eligible studies were identified from the hits. The overall pooled correlation of body mass index and blood pressure in the United Kingdom was 0.6, demonstrating that there is a correlation between the variables. From three of the studies, a correlation between body mass index (Kg/m2) and systolic blood pressure (mmHg) across the participants was noted (n = 27,322, SD: 21.4; r = 0.6, p>0.000).

**Conclusion:** Body mass index and blood pressure are strongly correlated in both the general population and tens of thousands of subgroups, suggesting that almost all demographic groups in the UK are affected by the growth in hypertension prevalence. In light of the estimates of the potential associations between body mass index, stroke, and ischemic heart disease based on the current pattern of treatment in this population, the UK and other nations going through a similar epidemiologic transition should be persuaded to address the rising prevalence of hypertension as a national priority.

## 1.0 Introduction

According to Di Angelantonio *et al*. (2016) [1], body mass index is a measure of body fat that is calculated as the ratio of a person’s weight to the square of their height, expressed in kilogrammes per square metre. Because the same limits can be used for both men and women, it is the metric that is most frequently used to determine if a population is overweight or obese. According to the Centers for Disease Control and Prevention (CDCP) (2022) [2], a person’s body mass index (BMI) is determined by dividing their weight in kilogrammes (or pounds) by the square of their height in metres (or feet). A high BMI might be a sign of significant body fat. Blood pressure (BP) is defined by Ogedegbe and Pickering (2010) [3] as the pressure created by flowing blood on blood vessel walls. The National Institutes of Health (2022) and the American Heart Association (AHA) both emphasised that a normal systolic and diastolic blood pressure is less than 120 mm Hg [4]. Body mass index and blood pressure are positively correlated; this link has important ramifications for nations like the United Kingdom (UK), where obesity and hypertension are on the increase.

According to a research by Neter et al. (2013) [5] in the UK, it is essential to have a better understanding of the link between BMI and blood pressure in order to determine the impact and develop mitigation measures for it. Body mass index is one of the most important public health issues in the globe [6]. When elevated (BMI 30.0), it is a substantial independent risk factor for chronic diseases including cardiovascular disease and diabetes mellitus and is associated with high rates of morbidity and mortality [6,7, 8, 9, 10]. Some studies suggest that up to 20% of adults in wealthy countries may have obesity-associated hypertension, which may be the cause of 78 and 65%, respectively, of essential hypertension in both men and women [6, 11]. In the UK, the prevalence of obesity presently ranges from 10% to 20% for men and 10% to 25% for women [12]. In the UK, the Centers for Disease Control and Prevention (CDCP) conducted a survey in 2002 that found that 31% of individuals were obese and 65% of people were overweight [13]. Furthermore, research show that nine (9) million people in the UK pass away from hypertension each year [14, 15]. Additionally, several studies have shown a connection between increasing blood pressure and weight increase [8, 16, 17]. Data from the 2004 National Health and Nutrition Examination Survey showed that 15.3% of lean persons and 42.5% of obese adults with a BMI of > 30 kg/m2 had hypertension, respectively [18]. 60% of hypertension is caused by an increase in adipose tissue storage, and obese people are 3.5 times more likely to get it [7].

Over the past 20 years, obesity incidence has significantly climbed throughout the United Kingdom and parts of Europe. Many emerging countries in Asia have closely followed these trends [5, 19, 20]. Between 1980 and the late 1990s, the prevalence of obesity climbed from 10% to 40% in the majority of British cities. Obesity rates in the UK are currently between 10% and 20% for males and 10% to 25% for women [21]. In England, 64% of adults were overweight or obese in 2020 [19]. Diabetes, cardiovascular disease, and various malignancies are just a few of the major diseases and fatalities that are linked to being overweight or obese [22]. Between 2000 and 2020, the percentage of adults who are overweight or obese has significantly increased [19]. Over time, there has also been an increase in the percentage of persons with a particularly high waist circumference, a symptom of central obesity [23].

Most of the pressure is brought on by the heart’s ability to pump blood via the circulatory system [3]. The worldwide average blood pressure, which is about 127/79 mmHg in men and 122/77 mmHg in women in the UK, has stayed almost unchanged since 1975, despite the fact that these average values mask wildly divergent regional trends [24]. Some factors seem to play an important role, such as consuming too many calories coupled with not exercising enough to burn out the excess calories, a combination that easily results in becoming overweight, making it necessary to understand the relationship between the two variables considered in this study [25, 26].

### Rationale/Justification

There is little research that establishes that Blood pressure and body mass index has a link, which could be a cause of health problems in people. A person with a BMI that is higher than what is deemed appropriate for their height to weight ratio is susceptible to having high blood pressure, which can lead to health concerns like type 2 diabetes, gallstones, respiratory problems, and some malignancies [1, 12, 22, 27]. In the United Kingdom, the majority of people have blood pressures that are higher than ideal but less than the usual cut-off for diagnosing high blood pressure, which is between 120/80 mmHg and 140/90 mmHg [22, 23, 28, 29]. If people continue to remain ignorant, ineffective techniques to deal with these difficulties will be adopted, resulting in ineffective results owing to a lack of awareness resulting from a lack of studies. The association between BMI and BP is positive across tens of thousands of people, according to a 2018 PubMed cross-sectional study that included 1.7 million adults (aged 35 to 80 years) from 141 primary health care sites (53 urban districts and 88 rural counties) from all 31 provinces in mainland China. The study looked at the heterogeneity in the association between BMI and BP across a wide range of subgroups of the Chinese population [30].

Healthcare providers in a clinical setting provide and inform decision-making in the management of conditions such as obesity and hypertension. A healthy BMI for women and men is between 18.5 and 24.9 worldwide [31–33]. According to research, BMI and blood pressure are both on the rise around the world, according to research, and epidemiological studies demonstrate a favourable association between the two, as previously said. Harsha and Bray (2008) [34] observed that the causes of overweight and obesity are multifaceted, which implies that clinicians need evidence from multiple correlates of health indices to substantiate their findings. In addition, based on the unavailability of evidence correlating blood pressure and body mass index in the United Kingdom, findings from this current systematic review will add to existing literature, therefore aiding future researchers and policymakers with evidence to strengthen their claim. However, with the availability of primary data from scoping results, it is vital for the researcher to provide sufficient evidence on the correlative study on blood pressure and body mass index in the United Kingdom.

### Aim/objectives

To provide evidence based on the correlative study of body mass index and blood pressure in the United is the overall objective of this review.

### 2.0 Methodology

**This review** provided evidence on the methodology chosen for the study, the data extraction method, understanding the concept of systematic reviews and the protocol through which this current study was carried out, the search strategy, and the inclusion and exclusion criteria of studies evaluating the correlation between body mass index and blood pressure among participants in the United Kingdom.

### Information Source/Database/Search Strategy

According to Kapetanakis *et al.* (2014) [35], it is imperative to note the source and database from which information is drawn in a systematic literature review. The databases and sources through which records and literature will be drawn from make it credible for the systematic review to be reproducible. Truncations, wildcards, proximities, and Boolean operators (“OR,”“AND,”“NOT”) were used to increase the sensitivity of the search for information in the literature review. Primary articles addressing the correlation of body mass index and blood pressure in the United Kingdom published in the form of original research articles were considered, to maximise strength and limit the potential for bias. Also, hand searching was performed on the reference list of some potential records to identify studies that were included in the synthesis.

### Database

The databases for the literature search for studies include the following;

- PubMed
- Web of Science
- PsycINFO
- Science Direct
- Cochrane Library

### Keywords in Search

((“Blood Pressure”) OR (“Systolic Pressure”) OR (“Arterial Pressure”) OR (“Diastolic Venous Pressure”) OR (“Pulmonary Wedge Pressure”) OR (“Diastolic Pressure”) OR (“Pulse Pressure”) OR (“Portal Pressure”) OR (Haemodynamics) OR (“Central venous Pressure”) OR (“Pulse”) OR (“Normal Blood Pressure”) NOT (“Hypotension” OR “Hypertension”) OR (BP) OR (“Blood Pressure [MESH]”) AND (“Body Mass Index”) OR (“BMI”) OR (“Body Weight”) OR (“Underweight”) OR (“Overweight”) OR (“Obese”) OR (“Body Mass Index [MESH]”) AND (Correlation) OR (Relationship) OR (Association) OR (“Correlation [MESH]”))

### Selection Criteria

In formulating the selection criteria of the study, the SPIDER (**S**ample, **P**henomenon of **I**nterest, Design, **E**valuation, and **R**esearch Type) was used. The participants for this study were individuals in the United Kingdom aged 18 years and above, and the study excluded HIV positive participants, hypertensive patients, and obese participants. Pregnant women and adolescents suffering from chronic conditions such as cancer, etc. were also not considered for selection. The study considered studies published from 2000–2022 and quantitative studies and mixed-method studies. Only published articles reporting the primary aim of the study in English were considered. The selection criteria for the study are stated in table 1 below.

**Table 1:** Selection Criteria.

| Variables | Inclusion Criteria | Exclusion Criteria |
| --- | --- | --- |
| **Sample/Participants** | Individuals in the United Kingdom aged 18 years and above | HIV positive participants, hypertensive patients, obese participants. Pregnant women and adolescents suffering from chronic conditions such as cancer etc. |
| **Phenomenon of Interest/Focus** | Correlation of body mass index and blood pressure | Studies focusing on hypertensive or hypotensive patients, studies that are addressing obesity and other disorders related to body mass index |
| **Design** | Quantitative studies and mixed method studies | Qualitative studies |
| **Research Type** | Cross sectional studies, randomized studies, cohort studies etc | Editorials, commentaries, studies with lack of clarity in design |
| **Study Duration** | 2000-2022 | Studies published earlier than 2000, studies without year duration |
| **Language** | English Language | Studies not published in English |
| **Publication Status** | Peer reviewed published articles reporting the primary aim of the study | Preprints, unpublished reviews, |

### Study Selection and Data Extraction

A standardised data extraction form was prepared by the researcher for the extraction and selection of articles or records from the databases. The Microsoft Excel software was used by the researcher with the help of two research assistants to extract and select articles. The de-duplication of records was performed using Revman software 5.2 after they were transferred from excel by the researcher. Studies that failed to meet the selection criteria of the study were excluded from the selection process. The overall body mass index, overall blood pressure, the year of publication, the author’s names, the study design and setting, the title of the publication, and the correlation coefficient were extracted from the primary studies during the selection process by the researcher and the research assistants. Disputes between the researcher and the research assistants were settled through discussion.

### Quality Assessment

The critical appraisal risk of bias tool was used in determining the quality assessment of the studies included in this systematic review. The methodology of the primary studies was judged using this critical appraisal risk of bias tool, and a score of 1 or 0 was assigned based on the items provided in the risk of bias tool. In the situation where there was a lack of clarity, the question mark (?) was used. At the end, the overall score for the quality assessment of the individual articles was classified into poor, fair, and good quality. Disagreements between the researcher and the research assistants were resolved through discussion during the quality assessment of the studies.

### Data Synthesis

Data synthesis in systematic review involves the method in which data from various sources is combined to provide a generalised answer [35]. For this current study, a narrative synthesis, which involves textualizing findings, was the adopted approach used in summarising the findings of this review. To represent the results of the study, tables and figures were used to depict the correlation between variables and the interpretation of the study.

## 3.0 Results

A total of seven hundred and seventy-eight articles were identified after searches were conducted on numerous databases such as PUBMED, Web of Science, Cochrane, and CINAHL. A total of three hundred and seventy-four (374) studies were presented for screening by abstract and title; studies that failed to meet the eligibility criteria were excluded. Duplicated articles found during the search and excluded from the analysis were excluded from this review, and records that did not correlate with BP or BMI or non-English written or published articles were also excluded to prevent bias. After screening of one hundred and ninety-nine studies for full text screening eligibility, seven studies reporting the primary aim of this study were included in the synthesis [25, 26, 27, 34, 36, 37, 38]. Illustrated in Figure 1 below, is the PICO framework of this study.

**Figure 1:**
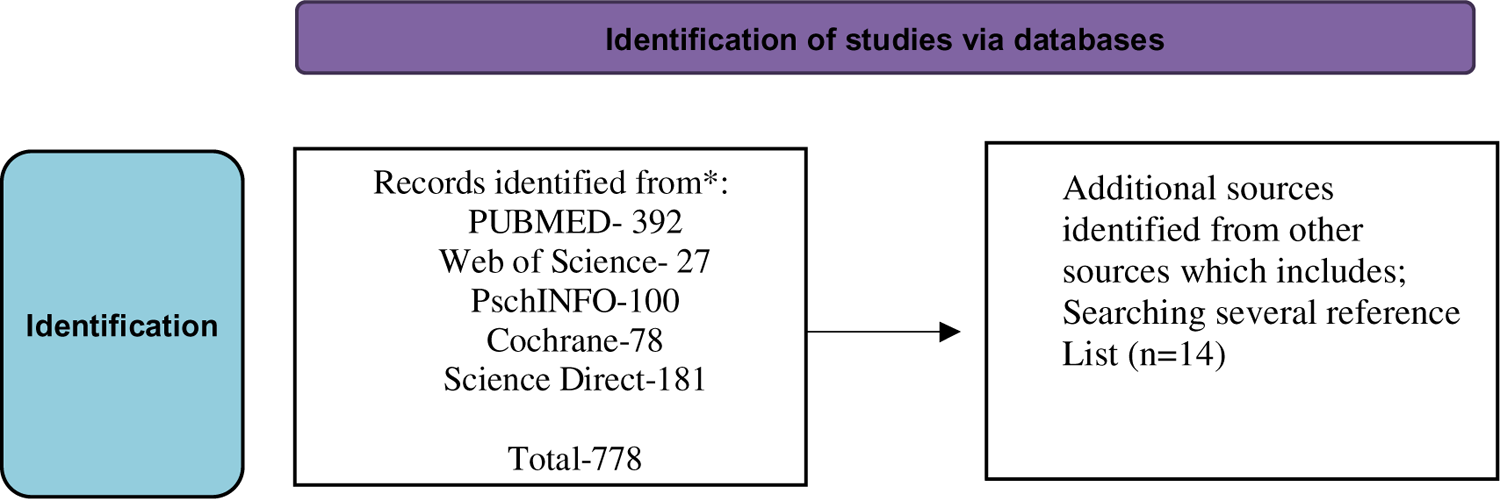
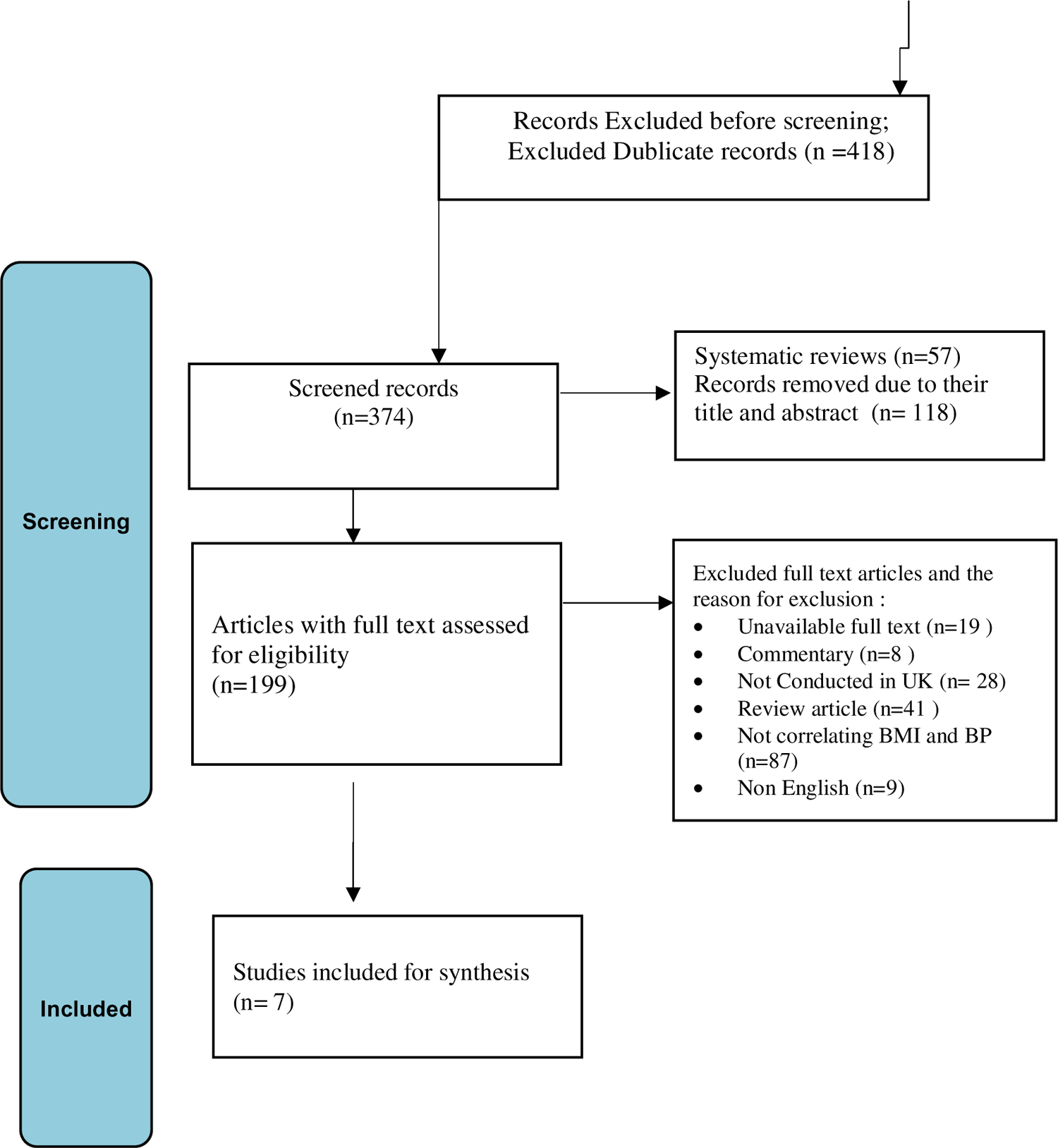
The PRISMA Diagram

### Characteristics of the Included Studies

A cumulative total of 27,322 participants were involved in the seven studies included in this synthesis [25, 26, 27, 34, 36, 37, 38]. Considering the design of the included studies in this review, three (3) of the included studies were cross-sectional studies [25, 34, 37]; two (2) studies were of cohort design [26, 27]; the review by Bernabe-Ortiz *et al.* (2021) [36] represented a pooled case series design; and one (1) other study was a comparative cross-sectional study [38]. Considering the setting in the United Kingdom where the studies were carried out, on the setting, two (2) studies were conducted in London [26, 37], one study in Northern Ireland [36], Leicester [34], two (2) studies in Wales [25, 27] and Yorkshire [38]. In seven of the studies, the average BP for diastolic and systolic pressure as well as the body mass index was noted in seven of the studies. Only six (6) studies showed the correlation between the NMI and BP [25–27, 34, 36–38]. The Table 2 below illustrates the included studies’ characteristics in thus synthesis.

**Table 2:** Characteristics of the Included Studies.

| Studies | Setting | Objective/title | Sample size | Study Design | Mean Blood Pressure (Systolic/Diastolic) mm/Hg* | Mean Body Mass Index (SD) (Kg/m<sup>2</sup>) | Correlation Coefficient (r) |
| --- | --- | --- | --- | --- | --- | --- | --- |
| Drøyvold *et al.* (2015) [26] | London, United Kingdom | The impact of the change in the body mass index on blood pressure | 2,524 | Cohort Study | 129.9/74.5 | 27.4 | 0.6 |
| Bernabe-Ortiz *et al.*, (2021) [36] | Newry, Northern Ireland | Correlation between blood pressure and body mass index levels a geographical setting and a cross sociodemographical: analysis of data pooled. | 21,902 | Pooled case Series | 125/73.4 | 25.4 | 0.5 |
| Harsha and Bray, (2008) [34] | Leicester | Weight loss and blood pressure control | 517 | Cross sectional Study | 126.2/78.8 | 25.9 | Not reported |
| Shuger *et al.* (2008) [25] | Cardiff, Wales | BMI used to predict hypertension incidences among normotensive women who were initially healthy | 98 | Cross sectional Study | 123.9/77.2 | 27.3 | 0.5 |
| Dua *et al.* (2014) [37] | London, England | Evaluation of BMI in relation to blood pressure among adults | 342 adults | Cross Sectional Study | 133/75.1 | 26.2 | 0.7 |
| Gelber *et al.* (2007) [27] | Wales District | A prospective study of BMI and risk factors, among men in developing hypertension | 1,826 | Cohort Study | 127/79.4 | 28.5 | 0.4 |
| Kamal *et al.* (2017) [38] | Yorkshire, England | Relationship between BMI and BP in Elderly English Men | 113 | Comparative Cross Sectional study | 125.2/75.8 | 25.9 | 0.5 |

### Quality Assessment and Critical Appraisal of Included Studies

Quality assessment of the included studies in the review on the association between blood pressure and body mass index is illustrated in Table 3 below. The results of these assessments showed that 4 of the studies had significant good quality [26, 27, 36, 38] and others had fair quality during the judgement for their methodology.

**Table 3:** Quality Assessment and Critical Appraisal of Included Studies.

| Statement | Drøyvold *et al.* (2015) [26] | Bernabe-Ortiz *et al.*, (2021) [36] | Harsha and Bray, (2008) [34] | Shuger *et al.* (2008) [25] | Dua *et al.* (2014) [37] | Gelber *et al.* (2007) [27] | Kamal *et al.* (2017) [38] |
| --- | --- | --- | --- | --- | --- | --- | --- |
| Is there a clear statement of the Objectives? | Yes | Yes | Yes | Yes | Yes | Yes | Yes |
| Is the research question viable for the research? | Yes | Yes | Not sure | Yes | Not sure | Yes | Yes |
| Are there clear procedures for justification of the study? | No | Yes | Yes | No | No | Yes | No |
| Is the methodological approach suitable for the study? | Yes | Yes | Yes | No | Yes | Not sure | Yes |
| Is the method of data collection viable based on the study methodology? | Not sure | Yes | No | Not sure | Not sure | Yes | Not sure |
| Is there a clear statistical procedure? | Yes | Yes | Yes | Not sure | Yes | Yes | Yes |
| Are there appropriate measurements in the study? | Yes | Yes | Yes | Yes | Yes | Yes | Yes |
| Is the method of data analysis suitable for the study research question? | Yes | Yes | Yes | Yes | Yes | Yes | Yes |
| Risk of bias stated? | Yes | No | No | Not sure | No | Yes | No |
| Are the outcomes of the review clearly stated according to the research questions? | Yes | Yes | Not sure | Yes | Yes | Yes | Yes |

### Concept of body mass index and blood pressure in the United Kingdom

This synthesis of seven research found that the ideas of body mass index and blood pressure [25–27, 34, 36–38]. The reviews by Dua et al. (2014) [37] and Gelber et al. (2007) [27] state in their text that blood pressure is one of the parameters that is most frequently monitored in clinics and that its readings are the main factors that determine which treatments are chosen. Four (4) research showed that arterial pressure is influenced by the heart’s output into the arteries, the flexibility of the artery walls, and the rate of blood flow through the circulatory system’s blood vessels [25, 27 37, 38]. Three forces kinetic energy, gravitational energy, and elastic energy affect a vessel’s pressure [25,38]. A thorough understanding of the connection between body mass index and blood pressure is crucial, claim Dua et al. (2014) [37]. Several of the included research have shown how convoluted the relationship is between hypertension and obesity, particularly how obesity-related hypertension is tightly connected with a number of other disorders as obesity progresses [34, 36, 38]).

Furthermore, there have lately been some concerns raised about the use of the most used anthropometric measure, BMI, as an accurate indication of obesity and body weight because it does not depict body fat distribution [36]. Additionally, its capacity to forecast the risk of cardiovascular illnesses and hypertension is questioned [27]. According to Kamal et al. (2017) [38], 66 to 79 percent of primary hypertension patients are related to obesity. They claim that having extra fat tissue might result in complicated bodily changes that aggravate or cause hypertension [38]. The sympathetic nervous system is overactive, adipose-derived cytokines (hormones) are altered, insulin resistance occurs, the kidneys’ structure and function are altered, and the renin-angiotensin-aldosterone (RAAS) system is stimulated. According to Kamal et *al*. (2017) [38], abnormal hormone signalling in obesity can lead to or exacerbate hypertension. Other ways that obesity exacerbates hypertension include changes in the function of the sympathetic nervous system, a part of the autonomic nervous system in control of the fight-or-flight response, and adjustments in the function and structure of the kidneys. More than 63% of all occurrences of cardiovascular disease are caused by blood pressure and BMI [25, 27, 37, 38]. Furthermore, more than one in four persons in England suffer from high blood pressure due to the high rate of obese and overweight people in the UK [25, 27, 36–38].

### Blood Pressure and Body Mass Index Correlation in the United Kingdom

Table 4 below illustrates the correlation between BMI and BP in the UK. The average blood pressure for diastolic and systolic pressure as well as the body mass index was noted in seven of the studies [25–27, 34, 36–38]. Only six (6) studies showed the values for the relationship between the blood pressure and the body mass index [36, 37, 38] [25–27]. The overall systolic blood pressure from the seven studies was 126.3. The overall pooled correlation of BMI and BP in the United Kingdom was 0.6, demonstrating that there is a correlation between the variables. From three of the studies, a correlation between body mass index (Kg/m^2^) and blood pressure (mmHg) across the participants was noted (n = 3623, SD: 21.4;r-0.6; p>0.000).

**Table 4:** Correlation of Blood Pressure and Body Mass Index in the United Kingdom.

| Studies | Average Systolic Blood Pressure (SD) mm/Hg* | Average Diastolic Blood Pressure (SD) mm/Hg* | Average Body Mass Index (SD) ($\text{Kg/m}^2$) | Correlation Coefficient (r) |
| --- | --- | --- | --- | --- |
| Drøyvold *et al.* (2015) [26] | 129.9 (18.4) | 74.5 (10.3) | 27.4 | 0.6 |
| Bernabe-Ortiz *et al.*, (2021) [36] | 125 (17.1) | 73.4 (12.2) | 25.4 | 0.5 |
| Harsha and Bray, (2008) [34] | 126.2 (16.4) | 78.8 (11.2) | 25.9 | Not reported |
| Shuger *et al.* (2008) [25] | 123.9 (17.0) | 77.2 (10.8) | 27.3 | 0.5 |
| Dua *et al.* (2014) [37] | 133 (18.2) | 75.1 (11.7) | 26.2 | 0.7 |
| Gelber *et al.* (2007) [27] | 127 (16.5) | 79.4 (12.2) | 28.5 | 0.4 |
| Kamal *et al.* (2017) [38] | 125.2 (17.4) | 75.8 (10.7) | 25.9 | 0.5 |
| **Overall Pooled Result** | **126.3** | **76.1** | **26.9** | **0.6** |

## 4.0 Discussion

This study provided evidence correlating BP and BMI in the United Kingdom among seven studies included in the synthesis. Based on the data and information from the records, it was determined that the research included in this synthesis on the correlation between blood pressure and body mass index in the United Kingdom involving adults showed a decreased link involving average blood pressure and body mass index [25–27, 34, 36–38]. This statement is consistent with studies assessing the effects of body mass index and blood pressure [39–41]. In four of the studies, further analysis revealed that in this correlation, there was no clear sign of a systematic trend, and in recurrent data from cross-sectional studies, a change was seen in the systematic process. Nevertheless, after synthesis and overall completion of the evidence, BMI remained significantly correlated with standard blood pressure, highlighting the long-term negative effects of the ongoing obesity epidemic [27].

It is necessary to explain the disparity in findings between the included research on the correlation of blood pressure and body mass index. The fact that previous research with a similar focus to this one has revealed increasingly strong cross-sectional body mass index-standard blood pressure relationships [42, 43, 44]. Additionally, as indicated in the seven investigations [25–27, 34, 36–38], the results of this synthesis may contain random discrepancies in findings as a result of sampling. Only women who were involved in three of the seven studies’ main observations were sampled. According to other research, unreported variations in the methods used to monitor BP and BMI may have also contributed to these variations [45, 46]. More measurement error when calculating BMI might diminish the relationships between BMI and standard blood pressure (due to regression dilution bias), but this explanation is implausible given that each cohort employed the same body mass index measurement procedures. Notably, each group employed a different set of blood pressure measuring tools. Even while the connection between systolic blood pressure and body mass index was shown to be smaller in studies by Harsha and Bray (2008) [34] and Bernabe-Ortiz *et al.* (2021) [36] (available data upon request), it can be conceived that the said calibration did not calibrate standard blood pressure instruments equally. In four studies that were part of this synthesis, the average difference in average BP per 1 kg/m3 rose at a later age. The observation that the BMI-diastolic BP correlations were weaker in some earlier research, which coincided with a similar trend in the present data, may potentially be explained by such differences. It is also clear and suggests that differential error has occurred from the current study on the association between body mass index and blood pressure (e.g., systematic underestimation of standard BP or diastolic BP among individuals with higher body mass index or overestimation among those with lower body mass index). To verify this hypothesis, studies including measurements of systolic blood pressure from numerous devices and measurements of anthropometrics are needed. This hypothesis has wider ramifications since it has the potential to skew comparisons conducted inside (longitudinally) and across studies.

The debatable present publication that reviewed trends in cross-sectional body mass index-blood pressure associations over time may be clarified by six of the seven studies included in this review [25–27, 34, 36, 37]. On the other hand, Kamal *et al.* (2017) [38] demonstrated that participants’ body mass index was impacted by complementing traits. The amplitude of these connections showed significant year-to-year variation, demonstrating how comparing them to two survey years may result in possible false findings about long-term trends. Some earlier studies [47, 48, 49] where an increase in strength of association was reported used hypertension as a possible outcome (which means, high BP or use of BP lowering treatments), and their outcomes may therefore be described by the trend of increasing treatment use (disproportionately rising hypertension prevalence among those at highest cardiovascular risk) [50]. The studies on the correlation between BMI and BP included in this synthesis demonstrated that continuous body mass index and BP measurements are interchangeable. Over time, weakening of the BMI and BP correlation has equally been observed in [27, 34, 37], while in some comparable reviews and some model specifications [49]. However, this review was limited to a “healthy sample” of 20–59 year-olds who did not have chronic prevalent conditions and who did not make use of long-term medications (antihypertensive included), and it indicated a growing connection [37]. In addition to this evaluation, quantile regression analysis was used to analyse the initial research. The outcomes show that the impact of BMI on BP seems to be obvious in both the blood pressure distributions above and below the threshold for hypertension treatment.

Both factors that can make the correlation over time weak (e.g., reduction in salt intake among individuals with a higher BMI)and those that could make it stronger (e.g., an increased fat mass for a certain BMI value in very recent decades) are likely to affect the magnitude of the relationship between BMI and underlying BP [51]. While some of the studies cited in this review state that the studies they used didn’t have time series data for these factors, the review outcomes of constant associations among younger-middle-aged adults from Harsha and Bray (2008) [34] and Bernabe-Ortiz *et al.* (2021) [36] suggest that these contradicting methods may have counterbalanced each other, which led to the same magnitudes of association seen in earlier studies [52–54]. The relative balance of these factors apparently resulted in a weakening of the association over time in older adults. While we do not have direct proof of an increase in the percentage of fat mass in the context of this study, proof from the United States and other places suggests that it could have occurred along with the increasing obesity epidemic [52]. In the UK, current public health initiatives have added blood pressure examinations for people who are most at risk for cardiovascular disease [45, 46, 52]. As a result, more people with an increased BMI are seen to be hypertensive and are receiving treatment for it [50]. While the prevalence of treatment has grown, as shown by our data, it appears to have had little impact on the correlations between BP and BMI, especially in younger age groups. The systematic review, which contends that the bulk of the body mass index-standard blood pressure relationship is seen below the 140 mmHg antihypertensive thresholds, serves as evidence for this. Modifications in health behaviours could have correspondingly had an impact on these processes. Wang et *al*. (2011) [51] claim that there has been a decline in salt intake in the UK recently, which may have helped certain people more than other individuals with increased BMI indices. BMI and BP may be further weakened by making further changes in the risk factor profile of individuals with increased body mass indices, like decreasing calorie and salt consumption and reducing sedentary lifestyles.

### Strengths and Limitations of the Review

Considering this study on the correlative evidence on body mass index and blood pressure in the United Kingdom, it is imperative to appraise the strengths and limitations of the findings of this synthesis. The use of cohort and cross-sectional research data improves the strength of this study and the evidence strength is that the sources of data where inferences were made are seen to be independent and also show complimentary attributes. The strength of this review is that potential co-founding roles of certain variables were neglected during the data extraction to make and robustly boost the strength of the evidence with a clear representation of the average standard deviation, diastolic blood pressure, systolic blood pressure, and correlation coefficient of the variables measured in the different studies of the review. Based on the limitations encountered in this study, being a student’s dissertation, it was difficult to involve credible independent reviewers to extract data and perform the quality assessment of the primary paper included in this synthesis. It has been reported that at least three impartial reviewers must participate for a systematic review process to be largely resistant to bias. Notably, missing data from the main studies was not taken into consideration, and many likely cofounders went unmeasured in several of the primary studies. Co-founding factors, such as demographic data and other socioeconomic characteristics, were not retrieved. The results of this study indicate that comparing association patterns before and after this year should not be done with caution because analytic weights were not introduced. Although it is difficult to compare research when BP measuring devices have improved, regression models based on studies concerning calibration were not employed in this study to take this into consideration, thus posing a limitation. Therefore, the validity of the presumptions behind such calibration is necessary for the interpretation of investigations. The study’s failure to add constant values to the raw diastolic and systolic blood pressure readings for individuals receiving therapy for hypertension is another weakness. A lack of information on therapy, dosage, and adherence makes it difficult to approximate the influence of treatment usage on relationships between BMI and BP, even though it was expected to have a negligible influence.

### Recommendations and Conclusion

From this study concerning the correlation between blood pressure and body mass index. The over-all population and tens of thousands of subgroups reveal a significant association between body mass index and blood pressure, indicating that the constant rising body mass index would be linked to the constant prevalence of hypertension among almost all demographic categories. In addition to other public health initiatives, treatment for high blood pressure modifies this link and may be essential in reducing the negative public health effects of rising BMI. Evaluations of the possible association between ischemic heart disease, stroke, and body mass index should compel the UK and other countries going through a similar epidemiologic transition to address the rising prevalence of hypertension as a matter of national priority. The consequences of body mass index may vary by age group and period. Age-group correlations between body mass index and standard blood pressure happen to have declined in recent decades, especially at older ages. The declining strength of the correlation may counterbalance the effect of the rising prevalence of obesity at the population level. Body mass index continues to be strongly correlated with standard blood pressure across all adults in the United Kingdom, emphasising the long-term negative effects of the obesity epidemic. Finally, these systematic review findings demonstrate the value of combining several datasets to derive reliable conclusions about changes in risk factor-outcome relationships over time, as well as the possible drawbacks of inferring long-standing health trends when assessing just two time points. A review has offered insight into the relationship or correlation between changing risk for hypertension and changing BMI levels. A cursory review of a few instruments revealed that body weight, erect height, blood sugar level, blood pressure, and total cholesterol in the blood can all be measured directly. It has been noticed that individuals are overweight in accordance with the WHO’s recommended body mass index cut-offs. Among individuals with an average BMI, the prevalence of hypertension summed up to 45%, when compared with 67% for overweight participants, 79% for participants in classes I and II for obesity, and about 87% for individuals in class III (p for trend 0.001). for obesity. Systolic and diastolic blood pressure distributions corrected for age were consistently and substantially varied based on body mass index level. The average diastolic and systolic blood pressure increased linearly and significantly across all body mass index levels. They asserted that body mass index, irrespective of other clinical risk factors, may have a different impact on blood pressure. This systematic review offered information on the correlation between body mass and blood pressure in the UK. The findings showed that there are several factors that affect how BMI and blood pressure are correlated. Corresponding with the systematic study, cardiovascular disease (CVD) and diabetes mellitus are the risk factors that influence this link. Also, it could be seen that there are no studies that have correlated blood pressure and body mass index of individuals in the United Kingdom, and as such, health practitioners in the field of nursing must consider the outcomes of this study in clinical practise and concerned organisations must build from the findings of this review. Further research is imperative, particularly with gaps in literature and future studies must be considered to explore other variables that are related to blood pressure and BMI, such as correlating conditions like hypertension and obesity, particularly among adults in the United Kingdom. It is also imperative that in nursing practice, considerations must be placed on adequate patient care and a clear understanding of the relationship must be used in delivering healthcare services to adults within the age group. Finally, doctors and nurses in practise will require data from a variety of correlates of health indices to support their conclusions. Additionally, because there is a lack of data linking BMI and blood pressure in the UK, the outcomes of this systematic review will contribute to the body of literature already in existence, providing future researchers and policymakers with more support for their claims. It is crucial for the researcher to offer enough support for the correlative study on BP and BMI in the UK nevertheless, given the availability of primary data from scope findings.

## Ethics Approval and consent to Participate

Not Applicable

## Consent to Publish

Not applicable

## Availability of Data and Materials

The Data set from the study are available to the corresponding author upon request.

## Data Availability

All data produced in the present study are available upon reasonable request to the authors.

## Acknowledgements

Not Applicable

